# Increased risk of death immediately after discharge from compulsory care for substance abuse

**DOI:** 10.1101/2022.01.16.22269380

**Authors:** Anders Ledberg, Therese Reitan

## Abstract

**Background:** In Sweden, approximately 1000 persons per year are committed to compulsory care for substance abuse for a maximum duration of six months. People admitted to compulsory care are known to suffer high mortality risks, but whether the risk of dying is further heightened immediately after discharge is not known.

**Methods:** Individual data from Swedish national registers were used to follow all persons discharged from a six months compulsory care episode in the period 2000–2017 (*N* = 7, 929). Based on a competing risks framework including re-admissions to compulsory care or imprisonment, hazard rates were estimated in five non-overlapping time windows covering the first year after discharge.

**Results:** In total, 494 persons died during follow-up, corresponding to an overall yearly risk of 6.2 percent (95% confidence interval: 5.7, 6.8). The risk was higher for men than for women and increased with age. The risk of dying during the first two weeks after discharge was higher than during the remaining follow-up period – hazard ratios comparing the first two weeks with subsequent time windows were between 2.6 (1.3, 5.0) and 3.7 (2.4, 5.9). This heightened risk in close proximity to discharge was only observed for deaths due to external causes, and only for people below the median age of 36 years.

**Conclusions:** The risk of dying immediately after discharge from compulsory care is very high, especially for younger clients, and more efforts should be made to prevent these deaths.

## 1 Introduction

Misuse of alcohol and drugs is associated with a substantially increased risk of morbidity and mortality (e.g., Ezzati et al. 2004), and most countries have consequently implemented measures aiming to reduce the health burdens of alcohol and drug use. In Sweden, the municipal social services are responsible for providing persons who misuse these substances with “help and care needed to get away from the misuse” (Social Services Act, SFS 2001:453). When a person risks severely harming his or her health, the well-being of his/her next of kin, or is about to cause irreparable damage to his/her future due to a particularly risky use of alcohol or illicit drugs, and this individual is furthermore not willing to receive care or treatment voluntarily, the social services are legally obliged, by the LVM Act (SFS 1988:870), to initiate an application process for coercive care for this person. The application is directed to an administrative court (Förvaltningsdomstol) that decides whether there are sufficient grounds for a commitment to coercive care (henceforth LVM-care). The maximum duration of a commitment to care according to the LVM Act is six months, after which the client must be discharged. The stated purpose of the compulsory care legislation is to motivate clients to enter care or treatment programs voluntarily and, by extension, to cease their riskful alcohol- or illicit drug use. Approximately 1,000 persons in Sweden (of which some 30 percent are women) are annually admitted to coercive care under the LVM Act. The National Board of Institutional Care (Statens institutionsstyrelse, SiS) – a government agency under the Ministry of Health and Social Welfare – is responsible for providing compulsory care and currently operates 11 residential institutions (LVM-homes) across the country. Previous research has shown that people discharged from LVM-care have a substantially increased risk of dying compared to the general population (Fugelstad et al. 1998, Gerdner 2004, Larsson & Leiniö 2012, Hall et al. 2015). For example, Fugelstad et al. followed 101 persons with a history of intravenous heroin use who were admitted to compulsory care in 1986-1988. They reported a crude mortality rate, over a minimum follow-up of five years, of 7.8 per 100 person years. This rate was the highest among all studies included in a large review of opioid related mortality (Degenhardt et al. 2011). Larsson & Leiniö followed all persons discharged from LVM-homes during 1999-2003, most of which were admitted according to the LVM Act. Within one year of discharge, 230 of 4314 clients had died, corresponding to a one-year mortality risk of 5.3 percent. The high mortality after LVM-care likely reflects the fact that compulsory care is only applicable to persons with a severe substance misuse in the first place.

The follow-up study of Larsson & Leiniö (2012) also showed that many clients have recurring commitments to compulsory care according to the LVM Act; 20 percent of their sample were re-admitted during the one-year follow-up. Furthermore, the rate of criminal convictions among LVM clients is high; according to Larsson & Leiniö, about one third of the clients were convicted for a crime in the year following discharge, and eight percent served time in prison. Since the risk of dying is considerably reduced while in institutional LVM-care or during incarceration, a failure to properly consider the client’s status after discharge is likely to entail an underestimation of the true risk of death outside residential care. Consequently, instead of assuming that the risk of death is uniform after discharge, a competing risks framework could be adopted to account for potential differences in mortality risks between the possible “states” after discharge.

These previous studies were also based on an implicit assumption that mortality risks are uniform over time. However, studies of people who have been released from prison show that the risk of death may be particularly increased during the first weeks after discharge (see Merrall et al. (2010) for review), and similar results have been observed also after discharge from substance abuse treatment (Ravndal & Amundsen 2010, Merrall et al. 2013, White et al. 2015, Maughan & Becker 2019). This increase in death rates in the time period immediately following discharge is often interpreted as partly being a consequence of reduced tolerance caused by a prolonged (and often involuntary) period of abstinence. Indeed, a study using Swedish data, found that the overall risk of death after discharge from imprisonment was particularly increased among people with a prior diagnosis indicative of substance use disorders (Chang et al. 2015). Taken together, these studies suggest that the risk of dying could be particularly high also immediately after discharge from compulsory care according to the LVM-Act. This is an important topic to investigate, both in order to evaluate the efficacy of LVM-care, and also to pin-point temporal contexts where more targeted interventions might be aimed in order to prevent preventable deaths.

The aim of this study is to examine mortality rates after LVM-care, with a specific focus on changes in mortality as a function of time after discharge. Using official administrative registers, all persons discharged after a six-month episode of LVM-care during 2000-2017, were followed with respect to: vital status, new episode of LVM-care, and imprisonment. A competing risks framework is used to handle the different possible outcomes after discharge from LVM-care. To test if mortality rates depended on time-since-discharge, time-dependent mortality rates were estimated using Poisson regression.

## 2 Data & Methods

### 2.1 Data

The data for this study were obtained by cross-linking administrative information provided by SiS, the agency providing for LVM-care, with data from the National Board of Health and Welfare (Socialstyrelsen, SoS), and The Swedish Prison and Probation Service (Kriminalvården, KRIM). Cross-linking was made possible by the personal identification number (PIN) assigned to all residents in Sweden. Entries in the registries that did not have valid PINs were excluded (34 cases). Another nine entries with reused PINs were also excluded from further analysis. Mortality data were obtained from the Cause of Death Registry (SoS), and the cause of death was categorized as an ‘external cause’ if the underlying cause of death, according to the registry, belonged to chapter XX of the ICD-10. Data on periods of imprisonment were obtained from the Prison Registry held by KRIM. The researchers only had access to anonymized data and the project was approved by the regional ethics committee in Stockholm (numb. 2017/2221-31/5).

#### 2.1.1 Selection of the study sample

The maximum duration of compulsory care according to the LVM Act is six months, and this study focuses on those who stay for this maximum duration. This group constitutes the majority of cases admitted to LVM-care. Most of the clients discharged before six months were, in fact, discharged within three weeks of admittance and likely represent a group with less severe alcohol- and drug use.

In more detail, the following three criteria had to be fulfilled in order to be included in the study sample: a court-ordered admission to compulsory care according to section 4 of the LVM Act, that at least 175 days had passed between admission and discharge, and that clients had been discharged “regularly”, i.e., not discharged directly to jail, prison, or hospital care, nor being deceased at discharge. Cases that met these criteria were identified by cross-linking all discharges between years 2000 and 2017 listed in the LVM-registry, held by The National Board of Health and Welfare, with the administrative data provided by SiS. Considerable care was taken to correctly classify the conditions at discharge, and to determine whether deaths occurring the same day as the discharge happened before or after the client actually was discharged.

### 2.2 Analysis

Clients were followed for up to one year following discharge from LVM-care. Many clients were committed to LVM-care more than once under the observation period, and in these cases, follow-up was from the first discharge only. Follow-up lasted for one year after discharge, or until the first of three competing events occurred: start of a new episode of LVM-care, start of an episode of imprisonment, or death. Deaths were further subdivided into external causes, and other causes.

A competing risks framework was adopted (Putter et al. 2007) and cause-specific survival models were fit to the data using survival package for R (Therneau 2021), and plots were made using the ggplot2 package (Wickham 2016). To test if mortality rates were dependent on time, the time period after discharge was divided into five non-overlapping time intervals with breaks at: 14, 31, 90, and 182 days. These break-times were chosen to have higher resolution close to discharge to better follow changes in mortality rates in close proximity to discharge. We note that the main results are not crucially dependent on these exact break-times (not shown). Time-interval-dependent hazard rates were estimated for external- and other causes of deaths separately using Poisson regression (e.g., Rodríguez 2007). Assuming that hazard rates are constant in each interval, the estimates from the Poisson regression can be interpreted as estimates of hazard rates (Laird & Olivier 1981, Rodríguez 2007). Calendar year of discharge, sex, and age were included as covariates in the regressions. For the plots of marginal probability, two age groups were used (above and below the median age), and in the regression analyses, five age groups were used. All computations were made in R (R Core Team 2021). Necessary data and R-code in order to reproduce the figures and tables is available here: github/aledberg/lvm.

## 3 Results

### 3.1 Sample overview

According to the LVM-registry there were in total 18,269 discharges from LVM institutions between years 2000 and 2017, corresponding to 11,580 unique individuals. Eleven thousand and thirty-five of these discharges fulfilled the inclusion criteria, corresponding to 7,935 unique individuals, 65 percent of which were male. Six persons died before the date of discharge, and these six cases were not included in the analysis. The remaining 7,929 clients constitute the study sample and from now on all results refer to this sample, unless stated otherwise. The average number of clients discharged per year was 440, and there were no obvious trends over the 18 years constituting the observation period (not shown). Supporting Fig 1 shows that the age distribution of the study sample ranged from 18 to 86 years and had a peak around 22 years. The median age at discharge was 36 years (37 years for men and 34 years for women).

**Figure 1:**
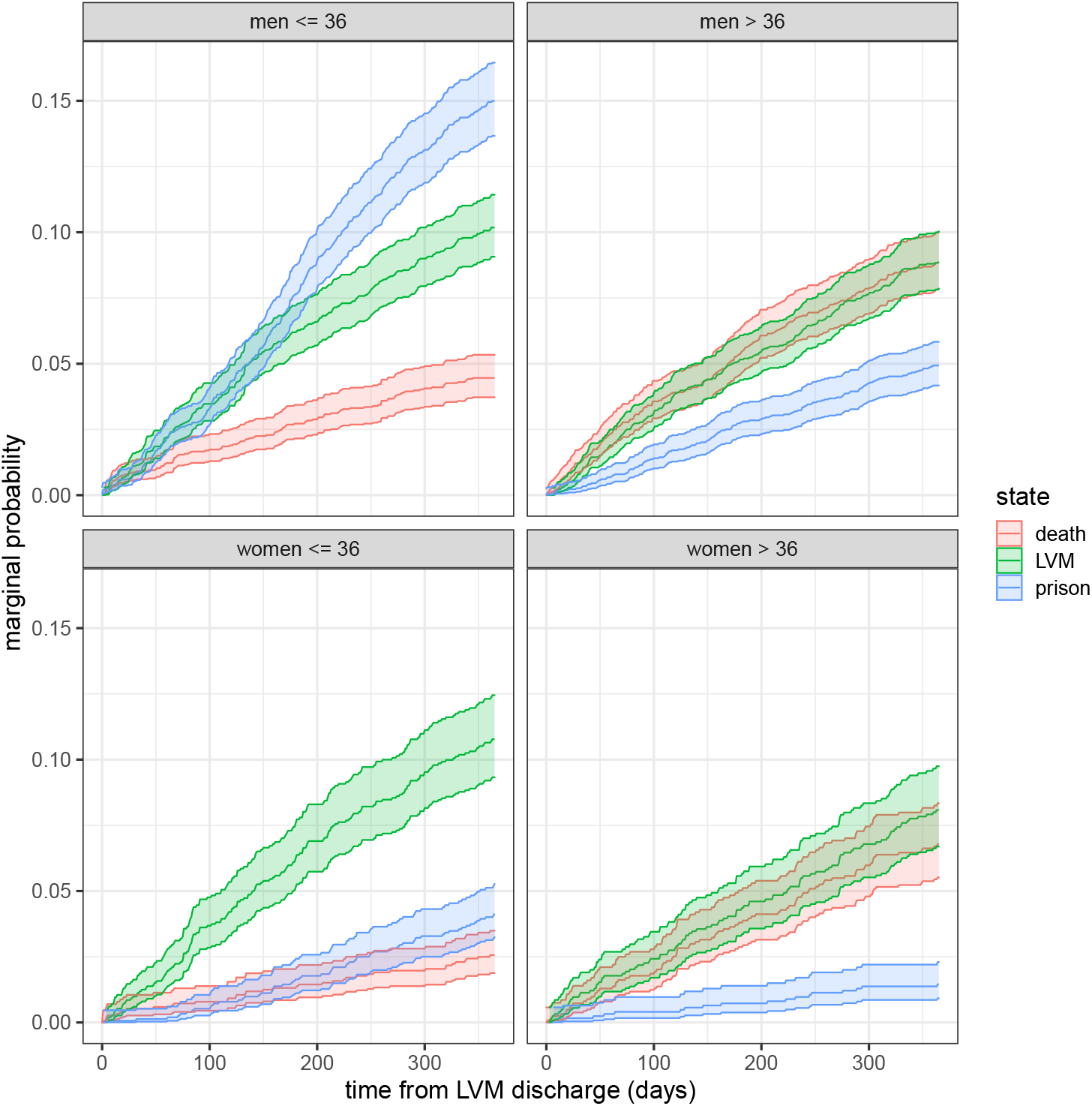
Probability of having entered into one of the three final states: new episode of LVM-care (LVM); imprisonment (prison), and death (death) by sex and age. Colored ribbons indicate 95 percent confidence intervals.

### 3.2 Overall survival

Within one year of discharge 494 people had died, corresponding to an annual mortality risk of 6.2 percent (95-% confidence interval: 5.7, 6.8). Among the deceased, 376 were male, mortality risk 7.1 percent, (6.4, 7.8), and 127 were female, mortality risk 4.6 percent (3.9, 5.5). Table 1 shows how mortality varied by age group and sex. As expected, the risk of death increased with age for both men and women. More than two thirds of all deaths in the youngest two age groups were declared to be due to external causes, the majority of which were results of poisoning (not shown in the table). The fraction of deaths due to external causes in the oldest two age groups were roughly one fifth.

**Table 1:** Number of persons deceased by age group and sex. **N**: number of persons discharged; **Deaths**: total number of deceased within a year from discharge, percentage classified as death due to external causes in parentheses; **Risk**: risk of death, (number of deceased divided by number of persons discharged), the 95-% confidence intervals are based on the Binomial distribution.

| Age | N |  | Deaths (% ext. causes) |  | Risk (95% C.I.) |  |
| --- | --- | --- | --- | --- | --- | --- |
|  | men | women | men | women | men | women |
| 18-25 | 1340 | 849 | 57 (77) | 25 (84) | 0.04 (0.032, 0.055) | 0.03 (0.019, 0.043) |
| 25-36 | 1194 | 673 | 61 (70) | 15 (93) | 0.05 (0.039, 0.065) | 0.02 (0.013, 0.036) |
| 36-50 | 1263 | 733 | 96 (46) | 31 (48) | 0.08 (0.062, 0.092) | 0.04 (0.029, 0.059) |
| 50-64 | 1116 | 416 | 122 (16) | 47 (23) | 0.11 (0.092, 0.129) | 0.11 (0.084, 0.147) |
| 64-86 | 256 | 89 | 31 (16) | 9 (22) | 0.12 (0.084, 0.167) | 0.10 (0.047, 0.183) |

### 3.3 Competing risks analysis

Clients were followed for up to one year after discharge from LVM-care or until they: i) started a new episode of LVM-care, ii) were imprisoned, or iii) died. A competing risk analysis was used to estimate the marginal probabilities of transitioning from the ‘discharged state’ into one of the other three states. Figure 1 shows the probabilities of ending in one of these states as a function of time in follow-up. This figure shows that men under 36 years of age (the median age) had the highest probability of entering into imprisonment; about 15 percent of the youngest males transitioned into this state within one year of discharge. The probability of being readmitted into LVM-care was similar for men and women and was slightly higher in the younger age groups. The probability of dying increased with age and was higher among men than among women, largely confirming the overall risks in Table 1.

#### 3.3.1 Cause-specific mortality

Causes of death were categorized as ‘external causes’ and ‘other causes’ and the probability of cause-specific mortality is shown in Figure 2. Observations were censored for anyone who first entered into prison or into a new episode of LVM-care. External causes of death dominated in the younger age groups whereas other causes were dominant among the clients 37 years or older. Furthermore, we note a steep rise in the probability of dying from external causes in the first few weeks after discharge in the younger age group. This will be examined more closely in the next section.

**Figure 2:**
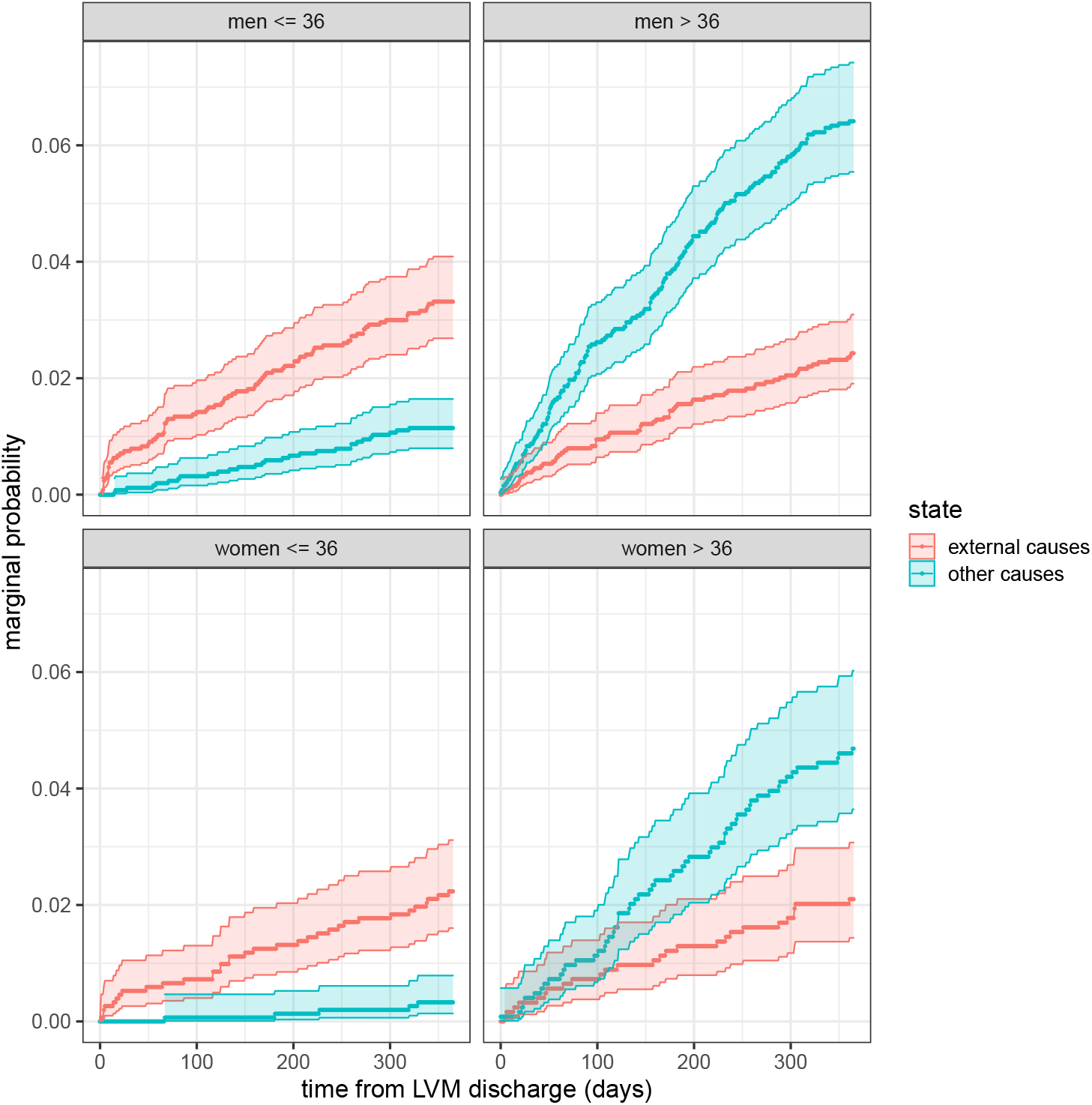
Probability of cause-specific mortality. Colored ribbons indicate 95 percent confidence intervals.

### 3.4 Time-dependent mortality rates

In order to investigate if the mortality rates depended on the time elapsed since discharge, the follow-up period was divided into five non-overlapping time intervals. Interval-specific hazard rates were estimated using Poisson regression for external- and other causes of deaths separately. The results for external causes are shown in Table 2 and for other causes in Table 3. For external causes of death (Table 2) the hazard rate was 2.6 times higher during the first two weeks after discharge compared to the following two weeks. An even larger difference was seen between the first two weeks and the final interval, i.e., the last half year of follow-up. A statistically significant relative increase in the hazards of deaths during the first two weeks were also observed when the analysis was repeated for men and women separately (not shown). However, in line with the results shown in Figure 2, the increase during the first two weeks were only readily observed in the youngest two age groups, i.e. among those below median age (not shown). Women had a reduced hazard rate compared to men and there was an increase in the hazard rate with year of discharge. The older age groups tended to have lower hazard rates compared to the youngest. For ‘other causes’ of death (Table 3) the hazard rates did not depend on the time since discharge. The hazard rate was lower for women compared to men but increased substantially with age for both sexes.

**Table 2:** Parameter estimates from the regression model for death due to ‘external causes’, by sex and age group.

| parameter | hazard rate | 95%-CI |  | p-value |
| --- | --- | --- | --- | --- |
| day 0-14 | 1 (ref.) | n.a. | n.a. | n.a. |
| day 14-31 | 0.39 | 0.20 | 0.75 | 0.0047 |
| day 31-90 | 0.30 | 0.18 | 0.50 | $2.4 \times 10^{-6}$ |
| day 90-182 | 0.32 | 0.20 | 0.50 | $9.4 \times 10^{-7}$ |
| day 182-366 | 0.27 | 0.17 | 0.41 | $2.1 \times 10^{-9}$ |
| year | 1.04 | 1.01 | 1.07 | 0.0035 |
| sex (women) | 0.70 | 0.52 | 0.94 | 0.02 |
| age 18-25 | 1 (ref.) | n.a. | n.a. | n.a. |
| age 25-36 | 0.98 | 0.68 | 1.41 | 0.91 |
| age 36-50 | 0.97 | 0.67 | 1.40 | 0.87 |
| age 50-64 | 0.62 | 0.39 | 0.96 | 0.03 |
| age 64-90 | 0.61 | 0.28 | 1.34 | 0.22 |

**Table 3:** Parameter estimates from the regression model using ‘other causes’ as the outcome.

| parameter | hazard rate | 95%-CI |  | p-value |
| --- | --- | --- | --- | --- |
| day 0-14 | 1 (ref.) | n.a. | n.a. | n.a. |
| day 14-31 | 1.15 | 0.57 | 2.36 | 0.69 |
| day 31-90 | 1.07 | 0.58 | 1.95 | 0.83 |
| day 90-182 | 0.85 | 0.47 | 1.54 | 0.60 |
| day 182-366 | 0.77 | 0.43 | 1.36 | 0.36 |
| year | 1.00 | 0.98 | 1.03 | 0.90 |
| sex (women) | 0.69 | 0.52 | 0.92 | 0.01 |
| age 18-25 | 1 (ref.) | n.a. | n.a. | n.a. |
| age 25-36 | 1.13 | 0.58 | 2.21 | 0.72 |
| age 36-50 | 4.00 | 2.34 | 6.84 | $3.8 \times 10^{-3}$ |
| age 50-64 | 10.22 | 6.16 | 16.97 | $< 2 \times 10^{-16}$ |
| age 64-90 | 10.83 | 6.00 | 19.53 | $2.5 \times 10^{-15}$ |

In order to further quantify the time-dependence of risk of death, the sample was restricted to those 36 years old or younger, and all-cause mortality rates were estimated for the first two weeks after discharge as well as for the remaining period. Estimates were made for men and women separately. The results shown in Table 4 show that the mortality rates were substantially increased during the first two weeks, both in relative and absolute terms. A mortality rate of 17 per 100 person years for young men is roughly 200 times the corresponding rate in the general population.

**Table 4:**
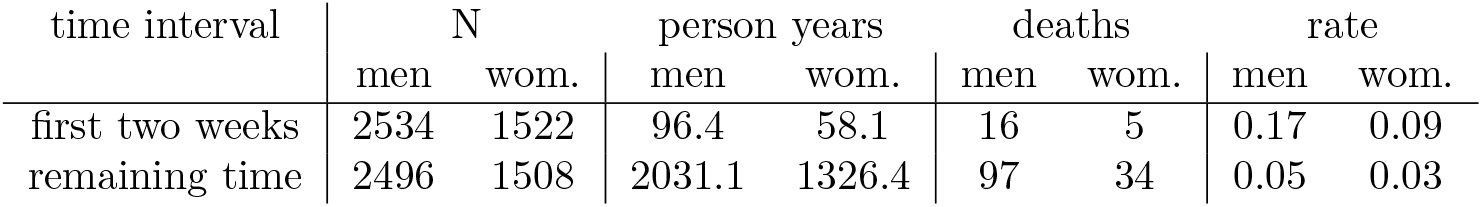
All-cause mortality rates as a function of time interval. Rates are estimated for those under 37 years of age and are expressed in units of per person year.

## 4 Discussion

### 4.1 Summary and interpretation of main findings

We examined the outcomes of clients discharged from compulsory care according to the LVM Act over the period 2000-2017, focusing on time-dependent changes in mortality. The overall risk of death during the one-year follow-up was 6.2 percent, in line with what has been reported by earlier studies (Fugelstad et al. 1998, Gerdner 2004, Larsson & Leiniö 2012, Hall et al. 2015). The risk was lower among women and younger persons (Table 1), but still substantial compared to the population at large as well as other clinical populations. For example, the mortality risk among the youngest age groups was higher than that observed among people undergoing methadone maintenance treatment in Stockholm during the same time period (Ledberg 2017). The risk of dying during the follow-up period was 11 percent among clients aged 50-64, more than 100 times higher than the corresponding risk in the Swedish population. Relatively few fatalities in this age group were due to external causes (< 20 percent), and the high mortality in this age group probably reflects the sequelae of long-term substance abuse.

To estimate the risk of death after discharge in further detail within this one-year follow-up period, a competing risk analysis was undertaken where imprisonment or a new episode of LVM-care were competing endpoints. This is the appropriate analysis given that one of the main aims of the study was to analyze time-since-discharge-dependent changes in mortality rates. Among men under 37 years of age, these competing endpoints were relatively common – about 25 percent were either imprisoned or were committed to a new episode of LVM-care during the one year follow-up (Figure 1) – demonstrating the relevance of the competing risk approach. To estimate changes in the mortality risk as a function of time since discharge, the follow-up period was split into five intervals and hazard rates were estimated for each interval using Poisson regression.

The analysis showed that the risk of death during the first two weeks after discharge was considerably higher than in subsequent intervals. In fact, hazard ratios were more than doubled (see Table 2). However, the increased risk during the first two weeks was only seen for external causes of death and mainly among younger people. Given that most of the external causes of death in the younger age groups consisted of poisonings, the most likely interpretation is that many clients return to the substance-use-habits they had before being committed to compulsory care, resulting in deaths from overdose for some. It is possible that a reduced tolerance following a period of forced abstinence contributed to the risk of fatal overdoses seen immediately after discharge. The relative difference in mortality risk between first two weeks and remaining time is similar to that reported in studies of people discharged from prison (e.g., Farrell & Marsden 2008, Bukten et al. 2017), and substance abuse treatment (e.g., Ravndal & Amundsen 2010, Merrall et al. 2013, Maughan & Becker 2019). In other words, the first few weeks after an abrupt increase in access to substances seem to be invariably associated with a heightened mortality.

In recent years it has become possible to receive opioid replacement therapy while still in LVM-care. This was not the case for clients in LVM-care during the time period analyzed in this study. It is therefore of great importance to investigate if this recent change in policy has had an impact on mortality rates overall as well as on rates directly after discharge.

### 4.2 Limitations

In the competing risk analysis we used three competing outcomes: death, a new episode of LVM-care, and imprisonment. This implies that we did not consider what happens to someone *after* they enter into a new episode of LVM-care or into imprisonment. An alternative would be to use proper multi-state modeling and estimate the risk of death from multiple states. For example, many clients have repeated entries into LVM-care and these repeated episodes could be modeled in future studies. In this study, analyses were restricted to the first episode for each person which implies suboptimal usage of data. However, given that the study sample included almost 8,000 individuals, and given the robustness of our findings, a relatively simpler analysis is, in our view, justified given the aims of the paper.

The five intervals used to investigate time-dependence of the mortality rates were chosen somewhat arbitrarily. Other studies have sometimes defined the first time-interval after discharge to be the first week (e.g, Farrell & Marsden 2008, Bukten et al. 2017) and sometimes the first four weeks (e.g, Ravndal & Amundsen 2010, Maughan & Becker 2019). These choices were likely dictated by the size of the available sample. Focusing on the first two weeks after discharge, as we did here, is in that way a pragmatic choice, but we note that the results remain qualitatively the same with a slightly shorter or longer first interval.

Even less guidance was available for cut-off points for the following intervals, and with the benefit of hindsight, the results would have been qualitatively the same with fewer intervals. In other words, for our purposes, the results did not depend on the exact details of the time intervals.

### 4.3 Deaths during admission

We have focused on the situation *after discharge* from LVM-care. However, it is highly relevant to also examine mortality risks *during* admission to LVM-care. Based on the data in this study, a client died while still in care in approximately 0.9 percent of the episodes of LVM-care. To properly interpret this mortality risk, it is necessary to examine the contexts surrounding these deaths. For example, it is not uncommon that clients abscond from LVM-care (Padyab et al. 2015), and it is therefore important to distinguish between clients who die while absent and clients who die while physically present at a LVM-facility. Unfortunately, the National Board of Institutional Care was unable to provide us with sufficiently detailed records to enable a closer analysis of whether the client in fact was present at the facility or died outside the premises.

### 4.4 Committed to care, but not necessarily confined

Compulsory care according to LVM means that a person can be withheld against his/her will in a confined facility for up to six months. However, the LVM-facilities consist of both locked and open wards providing clients with varying degrees of free movement within the general involuntary scheme. More importantly, though, the LVM legislation includes a “clause” (section 27) stating that all clients are, as soon as this is feasible, to be offered care outside the LVM facility by another provider than the National Board of Institutional Care. A closer description of this arrangement is provided by Reitan (2016). On average, around 75 percent of the clients are placed in care outside the LVM facility at least once during the period of commitment. It is, moreover, common that clients are placed in such alternative care at the time of discharge. That is, they are not physically present at the LVM facility although still under coercion. It is likely that clients who receive care outside of the LVM-homes constitute a less severe group with lower risk of death compared to clients who spend most of the period of commitment within the LVM facility. As already mentioned, given the data we were given access to, it was not possible to determine whether clients were actually present in residential care or not when he/she died. If we wish to find effective means of preventing post-discharge overdoses, we need to find predictors of increased risks. Systematic, reliable, and accessible data on clients’ whereabouts would enable us to monitor such risks in further detail.

### 4.5 Conclusions

We found that mortality after discharge from compulsory care for substance abuse was substantial, the one-year risk of dying was 7.1 percent for men and 4.6 for women, thereby confirming previous studies. Importantly, we also found that the risk of death due to external causes (mainly poisoning) were about three times higher during the first two weeks after discharge than during the remaining one-year follow-up. This novel finding indicates that more efforts should be made to prevent deaths in close proximity to discharge, especially for younger clients. These efforts may include both clinical and administrative measures, not least more detailed records about client movements within one episode of involuntary commitment.

## Data Availability

Data and code to reproduce the figures and tables are available online at

http://www.github.com/aledberg/lvm

## Acknowledgments

This research was funded by The Swedish National Board of Institutional Care (grant number: 2.6.1-1134-2017). The authors would like to thank Carl Pethrus at The Swedish National Board of Institutional Care for his assistance in determining the context of deaths that occurred in close proximity to the date of discharge.

## Notes

### Competing Interest Statement

The authors have declared no competing interest.

### Author Declarations

The researchers only had access to anonymized data and the project was approved by the regional ethics committee in Stockholm (2017/2221-31/5).

