## Supporting figure for "Increased risk of death immediately after discharge from compulsory care for substance abuse"

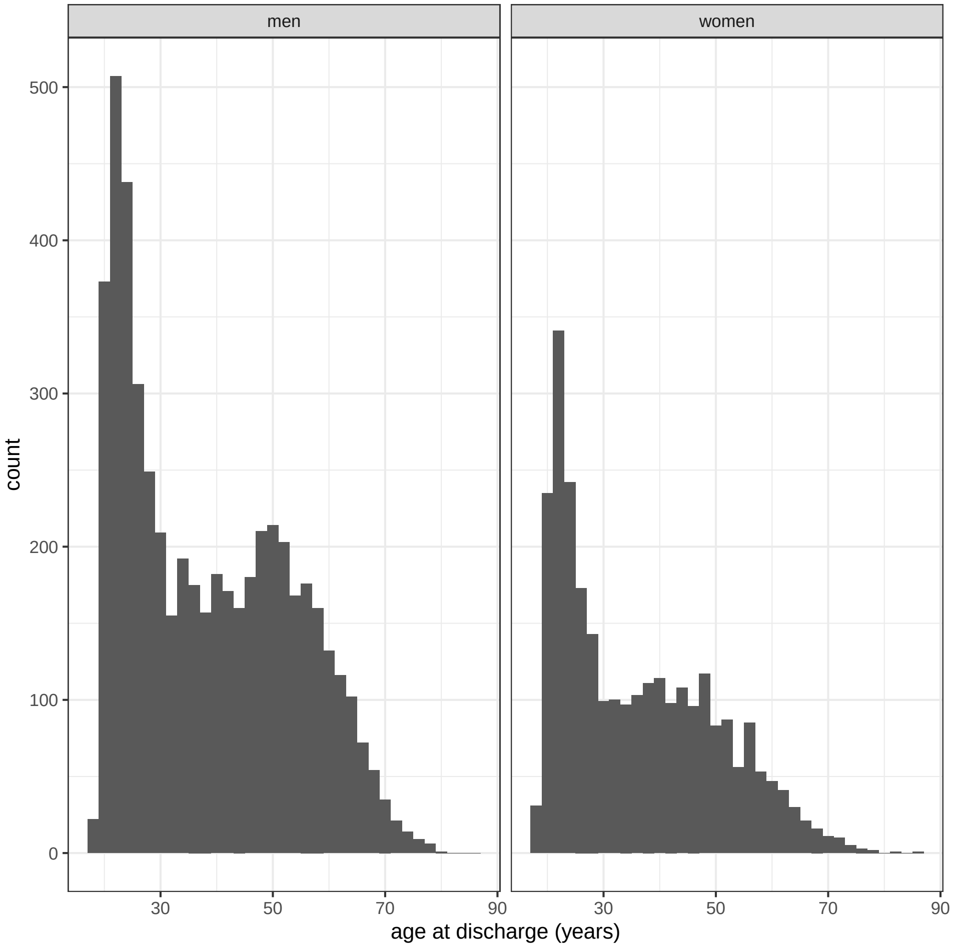


Figure S1: Age (in years) at discharge from LVM-treatment.
